# Middle Managers as Barriers or Enablers in Tackling Racial Discrimination in the NHS: A Qualitative Research Study

**DOI:** 10.1101/2025.02.27.25323018

**Authors:** Jaqui Long, Sandra Buchan, Fiona Sampson, Lilian Otaye-Ebede, Jeremy Dawson

## Abstract

**Objective:** To explore the role of senior and mid-level managers as barriers or enablers to change in tackling the discriminatory challenges experienced by Black and Minority Ethnic (BME) employees working in the NHS.

**Design:** A multi-level, multi-sourced qualitative study of five NHS Trusts in England.

**Setting and participants:** 26 qualitative interviews with senior leaders and BME network chairs (27 participants) and 5 focus groups (37 participants) with BME employees, across five NHS Trusts in England.

**Results:** Our findings revealed that discrimination, racial harassment, incivilities, lack of progression and exclusion experienced by BME employees, appear to be deeply ingrained in the culture of the NHS. Despite numerous national and local initiatives aimed to promote inclusivity and address discriminatory behaviours, our findings also revealed a notable disparity between what senior leaders thought was effective in addressing discriminatory behaviours and the actual lived experiences of BME employees. Finally, a key finding was the pivotal role middle managers played in setting the tone for whether discriminatory behaviours are challenged or allowed to persist, which directly impacts on the overall experiences of BME employees within the NHS.

**Conclusions:** Our results provide evidence that not only does racial discrimination continue to be experienced by NHS BME employees, but that middle managers are key to addressing and improving this situation. Despite there being national policies and initiatives addressing racial discrimination, our study found that positive change, whether at an individual or organisational level, is dependent on the actions and commitment of middle managers.

**STRENGTHS & LIMITATIONS:**

- Multi-sourced data from both senior leaders and BME staff
- BME staff in a variety of roles and grades
- Mixed qualitative method using interviews and focus groups
- Self-selected focus group participants may not be representative
- Participating Trusts actively motivated to address issues re: inequality – again less representative

## INTRODUCTION

The National Health Service (NHS) is the largest healthcare provider in the United Kingdom and one of the largest employers in the world (Ross et al, 2020), with employees represented from diverse backgrounds (Race disparity unit, 2019). Despite this diversity and the large numbers of Black and Minority Ethnic (BME) staff within the NHS workforce (Workforce Race Equality Standard – WRES, 2023), these staff continue to experience discrimination, racial harassment, abuse, incivilities and exclusion (Chastney et al., 2024; Adebowale & Rao, 2020). Specifically, studies have shown that ethnic minority employees are more likely to have negative work experiences, including being subject to stereotypes, biases and discrimination (Chastney et al., 2024). Similarly, the NHS Workforce Race Equality Standards (WRES) metrics demonstrate widespread race inequalities within NHS organisations, with a lack of inclusive cultures and predominantly white leadership, which does not represent the increasing racial diversity of the workforce (Pennington et al., 2023).

Pockets of research have created an awareness of these issues (e.g., Chastney et al., 2024; Kapadia et al, 2022), with some providing suggested actions (e.g. Pritchard, 2022). NHS England, the governing body of the NHS in England, is also aware of the ongoing issues, with many NHS organisations reporting a worsening trend (as detailed in the WRES 2023 report). As a response to these issues, the NHS has committed to tackling racial inequality at work and in healthcare through ongoing and established initiatives such as the Workforce Race Equality Standard (WRES), the NHS Race and Health Observatory and the NHS People Plan (NHS England, 2020). Reports from these initiatives have shown minimal progress and continue to call for more research into how we can create a fairer and more equitable workforce (WRES 2023 report, NHS EDI Improvement Plan, 2023). Therefore, urgent action and research is required to create in-depth understanding of the challenges and to identify solutions.

The role of middle managers in tackling race discrimination in the workplace continues to be unclear (Robinson et al, 2023). Our research aims to explore how leaders (senior and mid-level) are key to fostering or hindering progress made towards mitigating the negative persistent experiences of racial discrimination experienced by BME employees working in the NHS. We adopted a multi-perspective (i.e. BME staff and senior leaders) approach and sought to address the following research questions:

1. What are the key experiences (and resulting impact) of discrimination by BME employees in the NHS?
2. To what extent do leaders act as barriers or enablers to addressing these persistent race discrimination experiences at work?

## METHODS

### Theoretical Framework

We explore these research questions through the lens of ‘institutional theory’. Institutional theory provides a rich theoretical foundation for examining such critical issues such as race inequality within the UK healthcare sector. Underpinned by the notion of ‘institutions’ i.e., the “regulative, normative, and cognitive structures and activities that provide stability and meaning for social behaviour” (Scott, 1995, p. 33), we use institutional theory as a lens to examine how institutional structures, policies and practices systematically disadvantage certain racial groups (i.e., institutional racism). According to Jones (1997) the effects of institutional racism “are suffused throughout the culture via institutional structures, ideological beliefs and personal everyday actions of people in the culture” (Jones, 1997, p. 472). There are three levels within the organisation whereby institutional racism operates: the extra-organisational (i.e. between organisations and externals); the intra-organizational (i.e., the internal organisations climate, policies and procedures), and the individual (i.e., through employees’ attitudes, beliefs and behaviours) (Griffith et al., 2007).

### Setting

The data for this study was originally collected as part of a baseline analysis for a cultural change programme that the NHS England WRES team had planned to implement in 2020. Five NHS Trusts were identified and asked to participate based on their WRES indicators which highlighted areas for improvement in workplace culture for BME employees, however the programme was cancelled due to the COVID-19 pandemic.

### Ethical approval

The research was reviewed and approved by the Sheffield University Ethics Committee.

### Sampling, recruitment and data collection

The five NHS Trusts provided a diverse range of healthcare settings and geographical locations, adopting a comprehensive approach. Once agreement had been obtained, the national WRES team gave a presentation to senior staff to raise awareness and encourage participation.

Key informants for interview were identified and approached by the two researchers (JL and FS) and included the Chair of the Board, the CEO, one or more people with responsibility for HR and/or EDI and/or the WRES agenda, and the chair of the BME staff network. Participants were sent an information sheet and consent form and once the consent form had been returned, interviews were arranged. Interviews were conducted by one or two members of the research team and were undertaken predominantly via MS Teams, with a few being conducted by phone. This was due to the COVID-19 pandemic restrictions at the time. A topic guide was developed based on the stated objectives of the initiative and used to guide the interviews. Interviews lasted approximately 30-45 minutes and were recorded and transcribed, with the researchers also writing summary notes.

During the interviews with BME network chairs, an appropriate process for recruitment of BME staff to the focus groups was agreed. This varied between Trusts and included specific staff being directly approached by the BME chair; all staff in the BME network being emailed an invitation by the chair; and all BME staff being contacted via a list provided by HR. Potential participants were sent information and consent forms, which they returned to the research team who then arranged a convenient date and time for the focus group.

One online focus group for BME staff was held using MS Teams in each Trust. A topic guide was used to ensure key themes were explored, although this was used flexibly to allow participants to speak as freely as possible. Some people contributed via the chat function and in one case via a phone link due to connectivity difficulties. Focus groups generally lasted two hours and were video recorded and transcribed verbatim. The researchers also took notes and held brief reflective meetings afterwards to identify key issues.

### Analysis

We analysed the data using thematic analysis, exploring the personal experiences of discriminatory behaviour in the workplace and the strategies that effectively addressed them. Most of the analysis was undertaken by one of the researchers, with themes being checked with other members of the research team. Key themes were first identified across the senior leadership data for each Trust, and then across all five Trusts. A similar process was used to identify key themes in the focus group and BME chair interview for each Trust and then across the five Trusts. The two sets of summary findings were then integrated to identify the overarching key themes in the data.

### Positionality of researchers

Interviews were undertaken by FS and JL, with focus groups run by FS, JL and LOE. FS and JL are both health service researchers with no experience of working within the NHS and identify as white and female. LOE identifies as a black female with expertise in health service research but no experience of working within the NHS.

### PPI

None.

## RESULTS

We argue that configuration of the UK health sector, i.e., the NHS, provides a unique institutional framing heavily influenced by external regulatory bodies and a hierarchical management structure, that directly impacts on the norms that are embedded into the organisations culture/system and in turn individual attitudes and behaviours.

We undertook a total of 26 interviews, 21 with senior staff and five with BME network chairs, a total of 27 participants (one interview had two participants) (Table 1). We did not formally collect demographic data, but participants were approximately evenly split between men and women, and apart from the BME network chairs, were predominantly white.

**Table 1:** Interview Participants.

| Trust ID | Total participants | Role of participants (and number interviewed) |  |  |  |  |
| --- | --- | --- | --- | --- | --- | --- |
|  |  | CEO | Chair of Board | BME Network Chair | HR/Workforce/ Organisation Development Lead | EDI Lead |
| A* | 5 | 1 | 1 | 1 | 2 | 1 |
| B | 5 | 1 | 1 | 1 | 1 | 1 |
| C | 5 | 1 | 1 | 1 | 2 | - |
| D | 5 | 1 | 1 | 1 | 1 | 1 |
| E | 7 | 1 | 1 | 2 | 2 | 1 |

(\* One participant in this Trust had a dual role so total number adds up to more than 5)

We also undertook five focus groups with a total of 37 participants, (6-9 per group) (Table 2). Gender balance was approximately three quarters female. Not all participants provided data regarding ethnicity, but those that did described a range of backgrounds, with most describing Asian or British Asian, Black Caribbean or British Caribbean, and Black African, and a smaller number reporting mixed heritage. Participants were spread across a wide range of bands from 2 to 8d, plus one bank worker, with band 7 and above most strongly represented.

**Table 2:** Focus Group Participants.

| Focus Group Number | Trust ID | Number of participants | Researchers facilitating |
| --- | --- | --- | --- |
| 1 | A | 9 | FS, JL |
| 2 | B | 8 | FS, JL |
| 3 | C | 6 | FS, JL, LO |
| 4 | D | 8 | FS, JL, LO |
| 5 | E | 6 | FS, JL, LO |

We identified two key themes within the data which were present across all Trusts, despite their diverse contexts: experiences of discrimination at both an individual and structural level and the impact on BME employees; and the pivotal role of leaders and middle managers as barriers or enablers to change in tackling discriminatory behaviours.

### (1) Experiences of discrimination at individual and structural level and their impact on BME employees

Almost all BME staff described experiences of discrimination and disadvantage, both in terms of interactions with individuals and at a systemic or organisational level. They described greater levels of bullying and harassment, an issue also highlighted by senior staff from staff surveys. Whilst this sometimes took the form of overt racist and/or inappropriate comments, particularly in areas with a less ethnically diverse population, other more subtle forms of discrimination were also highlighted. Many described ‘micro-aggressions’, which left them feeling ‘unable to be their authentic self’ in the workplace. When these were challenged however, this could lead to negative reactions from colleagues, including blaming BME staff for taking things too seriously or being too sensitive, i.e. gaslighting. A lack of diversity in some teams could leave BME staff feeling isolated and over time this could undermine confidence and lead to self-doubt.

*“…some black staff, female black staff still feel that […] if they were to bring criticism or they were to voice unhappiness or something, they are perceived as the angry black woman. […] I’ve had Asian staff talk about when they’ve brought their lunch in and it’s been curry, for example, and the comments sometimes they get from non BME colleagues. […] being able to be your authentic self in the workplace has been difficult for some*.*” Trust B – BME chair*

In many instances, participants described a lack of support, engagement and/or recognition for the challenges they faced. This could include not challenging racist comments or behaviour from colleagues or service users, unwillingness to discuss issues relating to race, and expressing concerns about the fairness about initiatives to improve BME staff experience.

BME staff described feeling reluctant to come forward due to being labelled a troublemaker and the potential impact on their career. They expressed frustration and distress at the lack of response to sharing their experiences, raising concerns, or making suggestions, often at personal cost to themselves, and reflected how this led many people to stop engaging in consultations.

*“…some of my BME colleagues are fed up of even speaking up, because nothing changes even if people speak up” FGC P5*

*“Most of my colleagues here were really anxious to a point that some of the people were not willing to even take part, because they didn’t want to then be identified as the people who have, you know, erm, let the cat out of the bag*.*” Trust E BME Chair*

In addition to difficult interpersonal interactions, BME staff highlighted concerns regarding structural/systemic discrimination. One of the most frequently discussed issues was experiences of barriers to career progression. Many staff described their own personal struggles to progress, contrasting this with seeing their white colleagues advance more rapidly, despite having lesser skills, experience or qualifications. This impacted significantly on their confidence, making them less likely to apply for opportunities, and there were concerns that this could then be interpreted as a lack of motivation to progress.

*“…I have been that person in the last ten years, who’ve always been good enough to act but never actually get the role, and you actually always have your white colleagues getting the role but with less experience*.*” FGA P6*

*“I self-funded myself to get a master degree from [name] but still band 5 after working for over 17 years in the NHS. Same with all my BME colleagues*.*” FGC P4 (chat comment)*

Problems ranged from direct discrimination to a failure of managers to ‘go the extra mile’ to address existing inequalities. Many described repeatedly missing out on receiving information regarding development opportunities due to being in roles where they don’t have access to computers. When managers were challenged, it was brushed off and described as ‘a mistake’.

*“When you try to progress, everything you do has to go through online, but you know, some of us don’t have access to computer, we don’t work with computers, we work on the floor all the time*.*” FGA P9*

Other instances of discrimination included having applications for non-mandatory training repeatedly turned down, with a lack of transparency in decision-making processes. Concern was also raised about lack of access to informal coaching and information-sharing opportunities; these were linked to social networks which BME staff were not part of, and the need for formal processes to overcome this was highlighted. Even when successful in progressing, BME staff described experiencing suspicion from colleagues, with suggestions of positive discrimination and being a ‘token black’ rather than being appointed on merit. Others described occasional promotions as ‘tick box’ exercises to improve metrics, with a lack of support once in the role, and being judged more harshly for errors than white colleagues.

*“if you are promoted you’re either seen as the token black person and they’ve ticked a box. And then, to your peers, it’s ‘oh yeah you, you are that token black person, they’ve picked one and they’ve picked you*.*” FGB P5*

### (2) Role of Leaders as Barriers or Enablers

#### (2.1) The role of senior leaders as change catalysts

Whilst senior staff described being aware of the issues highlighted by BME employees, the focus group participants frequently reported lacking confidence in the leader’s commitment to address them. In some instances, the participants did not believe there was an understanding of the degree to which discrimination was experienced across the organisation.

*“…there’s a lot of sort of middle managers who don’t believe there is an issue, they don’t seem to, even with all the data there now*.*” FGB P3*

*“The willingness to change isn’t there with some of middle management. Some believe there isn’t an issue despite all the data out there*.*” FGB P6*

Senior staff described a range of initiatives to improve BME staff outcomes and experience, including reverse mentoring, inclusion of BME staff on interview panels, and talent management schemes. Whilst these were welcomed, many considered that they did not go far enough to address the underlying issues and achieve meaningful change, often only reaching those who were already engaged. Appointment of EDI posts at low grades in some instances also reinforced the sense that the issues were not being taken seriously. A particular area of concern highlighted by BME staff was the limited degree to which policies translated into action ‘on the ground’, and particularly a lack of engagement by middle managers to implement the policies.

*“…we may have all the flowery language and very good policies […] when it comes to crux of the matter, it’s the implementation” FGD P4*

*“There is no accountability. Ultimately what it comes down to is, there is no sufficient sanctions or accountability for managers’ actions*.*” FGB P1*

*“although you can speak freely and say your view and your point, it doesn’t always filter down. It’s almost as if some of the execs walk around with their eyes closed*.*” FGA P7*

*“There does feel like there’s the beginnings of a sense of change within the kind of executive leadership team within the Trust. So there seems to be a sense of commitment to wanting to create change, but (…) the layer that’s above me, so kind of my clinical leads, my service leads, the family service managers, are nowhere near that level of change*.*” FGD P5*

One positive recent change identified in most Trusts was greater support and engagement from senior leadership with the BME staff networks, including closer involvement in key decisions. Both COVID and the Black Lives Matter campaigns were seen as having catalysed this dialogue, and the tangible outcomes from it were leading to improved staff confidence and engagement with BME networks in most NHS Trusts, although the opposite was reported in one Trust.

*“Black Lives Matter has put us in a position whereby we’re able to have these conversations with our management. (…) for the first time in my life I had a corridor conversation with my manager about Black Lives Matter, and that’s when I got the confidence that oh is that a topic we can talk about now?” FGA/P6*

*“I’ve heard, you know, staff talking about “oh, you know, [P3]’s been to a BAME meeting again” FG/P3*

*“You just get pushed aside with those responses, “it’s the race card” oh, you know, it’s, you just become weary*.*” FGB/P8*

Many participants however highlighted the need for more resources to support the work of the BME networks, which frequently depends on participants’ and their managers’ goodwill. The need for more tangible commitment such as paid time or backfill of posts was seen as key to enabling more progress to be made.

#### (2.2) The pivotal role of middle managers in BME staff experience

Reflecting on the issues discussed in the previous theme, participants highlighted the significant impact their immediate line managers had on their workplace experience. ‘Middle managers’ were perceived as pivotal in either addressing race discrimination by actively implementing policies and providing direct support, or failing to do so by being insensitive and dismissive of BME staff experiences. In either case, managers were identified as the ones who set the tone for whether discriminatory behaviours were challenged or allowed to persist. Participants shared both positive, supportive relationships with managers and negative ones, recognising that these dynamics were crucial in whether discriminatory behaviours were challenged. The noted experiences ranged from highly supportive, to instances where managers failed to ‘go the extra mile’ to counteract discrimination, thereby reinforcing and compounding the situation.

*“I’ve worked under managers that have [a caring, nurturing, developing style], and it’s absolutely fantastic, I can tell you now. I don’t know if anyone else has, but it is really great, cause they’ll say well what do you want to be? I’m gonna help you – I’m gonna help you to get there*.*” FGC P4*

*“…my line managers are very, very supportive, I will say that*.*” FGC, P6*

*“…there’s too many times I’ve seen domestic and housekeeping staff being spoken to so disrespectfully, and a lot of managers think they can get away with it, you know, and it’s just, it’s just not right*.*” FGA P7*

*“The worst one I would say was*.. *when somebody had used the ‘N’ word in a meeting whilst I was at the toilet. Now, and I never, and my manager was in that meeting… I approached my manager and she’d not done anything about it*..*” FGB P8*

Due to disproportionately being at lower bands, BME staff often had white line managers, who in some instances did not appear to be comfortable or competent in building individual relationships with them. These and other experiences contributed to a lack of confidence to raise concerns.

*“…there seems to be a fear with some managers of speaking to BME staff, coz they don’t know how to approach us. Even though we all like food, we like sport*..*” FGB P3*

*“…stop calling me by somebody else’s name because they’re the only black person at the level, yeah? You know?! That’s not good. You know, you are my line manager, please get my name right…” FGA P1*

*“Staff experience goes probably unheard because your manager does not look like you, and you don’t feel you can necessarily trust them to share your real experience of the Trust*.*” FGA P3*

Many participants spoke of working harder to prove themselves and overcome discrimination, and feeling this was not recognised by their managers. Participants also reported not being understood or offered support when they raised concerns about unfair treatment or discrimination, or that no action was taken despite ‘saying the right things.’ In other instances, participants reported not being taken seriously, even with suggestions that they were exaggerating. Some described feeling blamed and labelled a ‘troublemaker’ and further discriminated against. These experiences unsurprisingly left BME staff unwilling to bring concerns to their managers.

*“…as well, you know, that the managers that say oh, I think there is an impression that we kind of overplay what we face on a day-to-day basis*.*” FGB P2*

*“If they think it’s ok to actually verbalise ‘oh, well I think it’s gone too far the other way now’ then, you know, how can they then help anybody progress or how can anybody go to them if they then think they’ve got an issue?” FGB P8*

*“…you always feel like you are a troublemaker…” FGE P1*

Participants recognised that many managers appeared unwilling, lacked insight or lacked the skills to discuss issues relating to race or to be challenged, with a fear of being seen as racist. Recommendations of additional training to address complex issues such as discrimination were regularly raised.

*“…what we need to try then is to try to change the attitudes and mindsets of managers so that they’re not just a manager for business, but they’re a manager for people to develop skills*.*” FGC P3*

## DISCUSSION

Our study offers one of the first qualitative multi-sourced studies exploring not just the persistent experiences of discrimination by BME staff in the NHS but goes further to reveal how and in what ways managers (particularly middle managers) act as enablers or barriers to addressing race discrimination issues at work. By addressing these research questions, we highlight how managerial actions impact BME staff experiences and the effectiveness of workplace initiatives.

Across all participants in our study, there was the recognition that BME staff experience needed to be significantly improved, both in relation to individual interactions with colleagues and through institutional level policies. Senior staff highlighted initiatives that were in place or being introduced to address these concerns, although there was variation in their extent and progression. Differing levels of understanding among senior staff of the issues relating to cultural change were also highlighted. In contrast BME staff views were much more consistent across all organisations. Whilst initiatives to bring about change were recognised, there was widespread frustration at the lack of progress. In particular, they frequently expressed concern about the ‘gap’ between the commitment being expressed at Exec/Board level and its implementation on the ground. The attitudes and behaviour of many middle managers was highlighted as a key barrier or enabler to change, and BME staff expressed varying levels of confidence in senior staff’s awareness of this, and their willingness to take action to address where barriers occurred. There was also widespread frustration that change was still being driven more by BME staff than the organisations that they worked for.

Whereas the importance of top-level or senior level managers roles in tackling race discrimination has been widely substantiated within literature (Cohen et al, 2013; Robinson et al, 2023), the role of middle managers remains an ambiguous topic (Robinson et al, 2023). This ambiguity has been suggested to be due to the challenges of having a dual role – one that is required to align with senior leaders as well as build trust within their teams (Floyd & Wooldridge, 1997; Huy, 2002; Mantere, 2008). This dual purpose creates potential conflicts, as middle managers must navigate pressures from above and below, making it difficult to balance the requirements placed upon them. It is the role of middle managers to embed organisational policies and strategies into operational priorities, however studies have shown that they may purposefully hinder organisational change (Gatenby et al, 2015; Kras et al, 2017). This suggests that their role in implementing policies, including those addressing discriminatory behaviours is not only complex, but may also involve resistance. This has been shown to be due to either a personal disagreement with the policy or the inability to manage the tensions it may create within their teams (Gatenby et al, 2015).

This study built on those that have looked at similar issues previously (e.g. Chastney et al., 2024; Linton, 2020) by involving both senior leaders and BME staff in a series of interviews and focus groups across five organisations. In this way we were able to identify the disconnect between the usually well-intentioned practices of senior managers, who were generally aware of issues and spoke of wanting to put them right, and the experience of staff at lower levels, where there was often little evidence of practices and policies making a difference to their working lives. In particular the pivotal role of middle managers of being the conduit for delivering better experience came through strongly. In contrast, a limitation of the study is that it included only five self-selecting organisations, which therefore might be those that are more aware of issues affecting BME employees. Future research may focus on a wider range of organisations, including those where senior managers are less alert to such problems.

A clear implication of this research is that senior managers need to ensure their actions are implemented at all levels, rather than assuming that policy changes at the top will necessarily impact the lives of those throughout organisations. Listening directly to those working in different roles, especially from BME backgrounds, can play a crucial role in that process. Likewise, it is important that middle managers are trained and supported to provide appropriate leadership and management which recognises and addresses the challenges faced by BME staff.

A limitation of this study is the potential impact of the researcher’s racial identities in facilitating the focus groups and interviews. The researchers who collected the data comprised of two white females and one black female, which may have influenced participant’s responses. While we acknowledged our racial identities at the beginning of each discussion, it is important to recognise that researcher positionality can shape both the dynamics of the conversations and the data collected.

## Data Availability

All data produced in the present study are available upon reasonable request to the authors

## AUTHORS CONTRIBUTIONS

Project design: JD. Data collection: FS, JL and LO. Data analysis: JL and FS. Drafting and reviewing paper: JL, SB, FS, LO and JD.

## ACKNOWLEDGEMENTS

We would like to thank all those who took part in interviews and focus groups for giving up their time and sharing their experiences.

